# Predicting Age-Related Macular Degeneration Progression with Contrastive Attention and Time-Aware LSTM

**DOI:** 10.1101/2022.05.19.22275305

**Authors:** Changchang Yin, Sayoko E. Moroi, Ping Zhang

**Affiliations:** The Ohio State University, Columbus, OH; The Ohio State University Wexner, Medical Center, Columbus, OH

**Keywords:** Disease progression prediction, patient subtyping, color fundus photography

## Abstract

Age-related macular degeneration (AMD) is the leading cause of irreversible blindness in developed countries. Identifying patients at high risk of progression to late AMD, the sight-threatening stage, is critical for clinical actions, including medical interventions and timely monitoring. Recently, deep-learning-based models have been developed and achieved superior performance for late AMD prediction. However, most existing methods are limited to the color fundus photography (CFP) from the last ophthalmic visit and do not include the longitudinal CFP history and AMD progression during the previous years’ visits. Patients in different AMD subphenotypes might have various speeds of progression in different stages of AMD disease. Capturing the progression information during the previous years’ visits might be useful for the prediction of AMD progression. In this work, we propose a **C**ontrastive-**A**ttention-based **T**ime-aware **L**ong **S**hort-**T**erm **M**emory network (**CAT-LSTM**) to predict AMD progression. First, we adopt a convolutional neural network (CNN) model with a contrastive attention module (CA) to extract abnormal features from CFPs. Then we utilize a time-aware LSTM (T-LSTM) to model the patients’ history and consider the AMD progression information. The combination of disease progression, genotype information, demographics, and CFP features are sent to T-LSTM. Moreover, we leverage an auto-encoder to represent temporal CFP sequences as fixed-size vectors and adopt k-means to cluster them into subphenotypes. We evaluate the proposed model based on real-world datasets, and the results show that the proposed model could achieve 0.925 on area under the receiver operating characteristic (AUROC) for 5-year late-AMD prediction and outperforms the state-of-the-art methods by more than 3%, which demonstrates the effectiveness of the proposed CAT-LSTM. After analyzing patient representation learned by an auto-encoder, we identify 3 novel subphenotypes of AMD patients with different characteristics and progression rates to late AMD, paving the way for improved personalization of AMD management. The code of CAT-LSTM can be found at GitHub^1^.

**CCS CONCEPTS:** • **Applied computing** → **Health informatics;** • **Computing methodologies** → *Neural networks*.

**ACM Reference Format:** Changchang Yin, Sayoko E. Moroi, and Ping Zhang. 2018. Predicting Age-Related Macular Degeneration Progression with Contrastive Attention and Time-Aware LSTM. In *Proceedings of SIGKDD 2022*. ACM, New York, NY, USA, 11 pages. https://doi.org/10.1145/1122445.1122456

## 1 INTRODUCTION

Age-related macular degeneration (AMD) is the leading cause of irreversible blindness in developed countries [20]. The number of people with AMD worldwide is projected to be 196 million in 2020, increasing substantially to 288 million in 2040 [22]. Based on clinical features, the disease is classified into early AMD, intermediate AMD (iAMD), and late AMD stages [12]. Late AMD is often associated with severe vision loss. Identifying patients at high risk of progression to late AMD, the sight-threatening stage, is critical for clinical actions, including medical interventions and timely monitoring.

Color fundus photography (CFP) is the most widespread and accessible retinal imaging modality; it is the most highly validated imaging modality for the detection of late AMD and the prediction of progression to the late stage of the disease [11]. Figure 1 show the CFP images of AMD progression for a patient’s eye. Some followup visits (e.g., the third year’s visit in Figure 1) might be missed. It takes 5 years for the eye to progress from early AMD stage to late AMD stage. Early identification of the risk of progression to late AMD and proper timely medical intervention might be able to alleviate disease progression.

**Figure 1:**
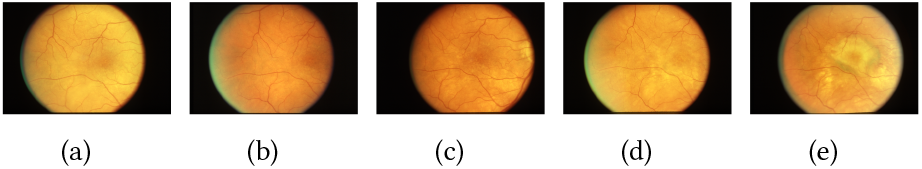
**Sample of AMD progression for a patient’s eye. (a) First year, early AMD. (b) Second year, early AMD; (c) Fourth year, iAMD. (d) Fifth year, iAMD. (e) Sixth year, late AMD. The third year’s follow-up visit is missed**.

Recently, deep learning methods have been proposed to detect abnormalities and AMD [4, 5, 7, 14, 18], and predict AMD progression [1, 3, 19, 23] in coming years based on previous CFPs. Although the above methods have achieved superior performance, they are limited to CFPs in the last visit and do not include the longitudinal CFP history and AMD progression during the previous years’ visits. The disease progression information can be found from the previous visits (as Figure 1 shows, patients usually have several visits before progression to late AMD). Patients in different subphenotypes might have various AMD progression speeds in earlier and later AMD stages. Capturing progression information during the last years’ visits might be useful for the prediction of AMD progression.

In this study, we propose a novel late-AMD prediction framework (**C**ontrastive-**A**ttention-based **T**ime-aware **L**ong **S**hort-**T**erm **M**emory network, **CAT-LSTM**) to model CFP sequences with consideration of AMD progression information. We utilize a time-aware long short-term memory (T-LSTM) to model patients’ temporal visits with irregular time gaps. The input of T-LSTM consists of genotype information, sociodemographics, CFP feature vectors and AMD progression information. Following [19, 23], we use the genetic risk score of 52 independent genetic markers as the genotype information, which has been reported to have associations with AMD risk in a recent large-scale genome-wide association study by the International AMD Genomics Consortium [13]. Following [3], we extract the smoking history, sex, age, race, body mass index as sociodemographic information. For the CFP features, we adopt DenseNet [16] to extract image feature vectors from CFPs. We introduce a contrastive attention module (CA) to remove the common features in the fundus image and learn fair image representation. We represent patients’ AMD stages during the last years’ visits as progression feature vectors. The concatenation of these four kinds of feature vectors is sent to T-LSTM to generate output vectors. Finally, fully connected layers and a Sigmoid function are followed to generate late AMD probability. Additionally, based on the learned fixed-size representations of temporal CFP sequences, we adopt k-means to cluster them into various AMD subphenotypes.

To demonstrate the effectiveness of the proposed framework, we conduct experiments on publicly available a real-world dataset from Age-Related Eye Disease Study (AREDS) [21]. The experimental results show that the proposed models outperform the state-of-theart methods.

In sum, our contributions are as follows:

- We develop a new LSTM-based AMD progression prediction framework CAT-LSTM that can model CFP sequences with irregular time gaps.
- We present a progression embedding module that can represent AMD disease progression information as vectors, which is helpful for late AMD progression prediction.
- We introduce a contrastive attention module that can force the model to focus on abnormal areas in CFPs by removing the common features in patient groups.
- We adopt k-means to identify AMD subphenotypes on temporal CFP data based on the well-learned CFP sequence representations.
- We demonstrate the effectiveness of our methods experimentally on real-world CFP data. By using T-LSTM and considering AMD progression information, our model outperforms the state-of-the-art AMD progression prediction methods by more than 3% on AUROC. Our subtyping framework identifies 3 novel AMD subphenotypes with different characteristics and progression speeds.

The rest of the paper is organized as follows. In Section 2, we describe our model in detail. In Section 3, we conduct experiments on real-world CFP datasets AREDS. We review the related studies in Section 4. Section 5 concludes our work.

## 2 METHOD

In this section, we propose a contrastive-attention-based time-aware LSTM (CAT-LSTM) to predict AMD progression and cluster CFP sequences into subphenotypes. We first present a CNN model with a contrastive attention module (CA) to capture abnormality from CFPs, and time-aware LSTM (T-LSTM) to model the CFP sequences and predict late AMD risks in coming years. Then we cluster the learned CFP sequence representations into subphenotypes with k-means.

### 2.1 Basic Notations

In this work, each patient’s data consist of a sequence of visits, which include CFPs for both eyes and sociodemographics. The elapsed time between successive visits are irregular. We treat the two eyes of a same patient as independent samples. Given an individual eye of a patient, the CFPs are represented as *V* = {*v*_1_, *v*_2_, …, *v*_*T*_}, where *T* denotes the number of visits for the patient. There are 9 steps (i.e., 1-9) for early and intermediate AMD stages, and 3 steps (i.e., 10-12) for late AMD stages [10]. The ground truth for the late AMD prediction tasks is represented as *Ŷ* = {*ŷ*_1_, *ŷ*_2_, …, *ŷ*_*T*_}, where *ŷ*_*t*_ ∈ {0, 1}. *ŷ*_*t*_ = 1 (*ŷ*_*t*_ = 0) denotes the corresponding eye will (not) progress to late AMD stage in coming years. We set different prediction windows for the late AMD prediction tasks. The sociodemographics of the patient are represented as *D* = {*d*_1_, *d*_2_, …, *d*_*T*_} ∈ *R*^*T*×*m*^. This work aims to detect abnormalities, predict AMD progression from CFPs. The framework of the proposed CAT-LSTM is shown in Figure 2. Based on the features extracted by CAT-LSTM, we further study the AMD subphenotypes with a subtyping framework as Figure 3 shown. We list the important notations in Table 6.

**Figure 2:**
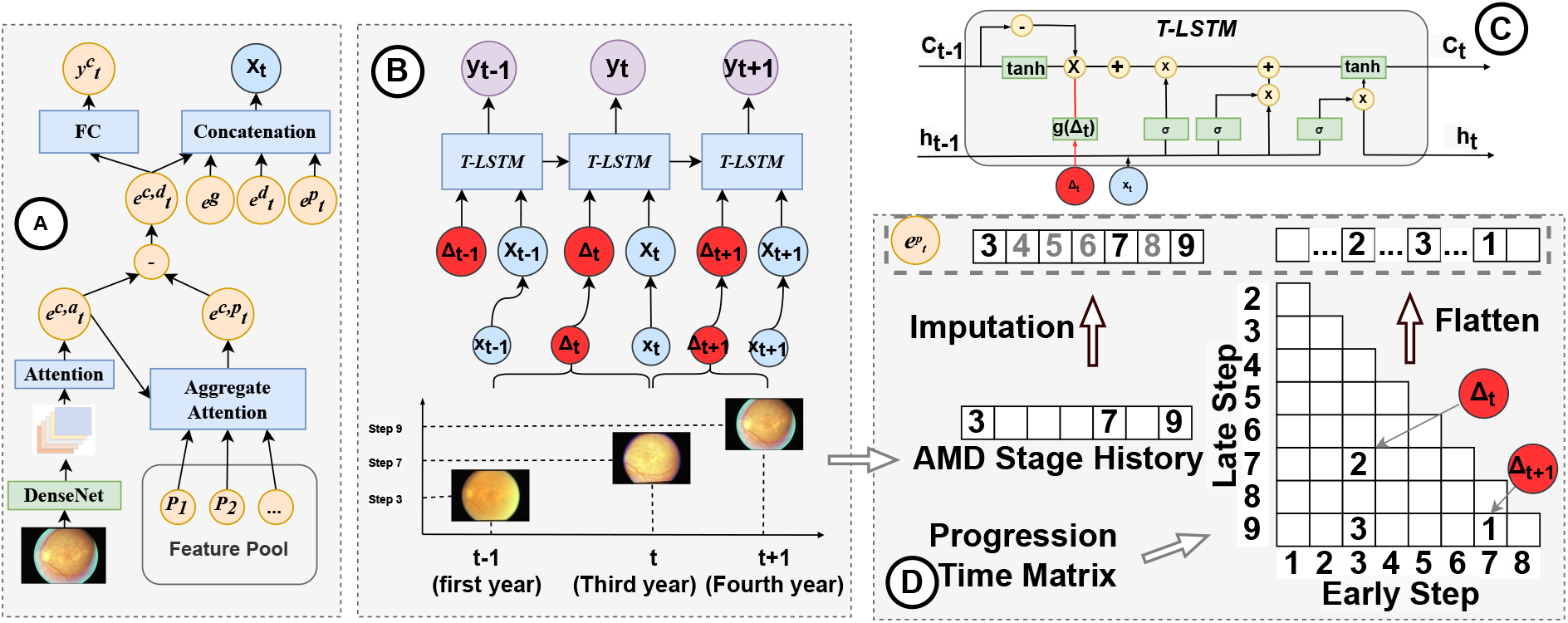
**Framework of CAT-LSTM. (A) CNN with contrastive attention module. we adopt DenseNet and an attention module to represent an individual eye image as a vector** 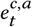. **Contrastive attention module removes the common feature** 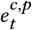 **and generate an abnormal feature vector** 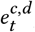, **which is used to predict AMD steps** 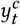 **in current visit. (B) T-LSTM. We obtain** *t*^*th*^ **visit’s feature** *x*_*t*_ **by combining the CFP feature** 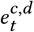, **genotype feature** *e*^*g*^, **sociodemographic feature** 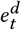 **and progression feature** 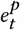. **Then** *x*_*t*_ **and time gaps** Δ_*t*_ **between successive visits are sent to T-LSTM to model the temporal CFP sequences. T-LSTM output the AMD prediction results** *y*_*t*_ **at each time step. (C) Details of T-LSTM. (D) Progression embedding. We generate an AMD stage history vector based on previous visits and impute the missing AMD steps. Then we build a AMD progression time matrix to represent how many years it takes for the eye to progress from earlier AMD step to later AMD step. We flatten the matrix and concatenate it with the imputed AMD step history vector to generate the progression embedding** 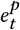.

**Figure 3:**
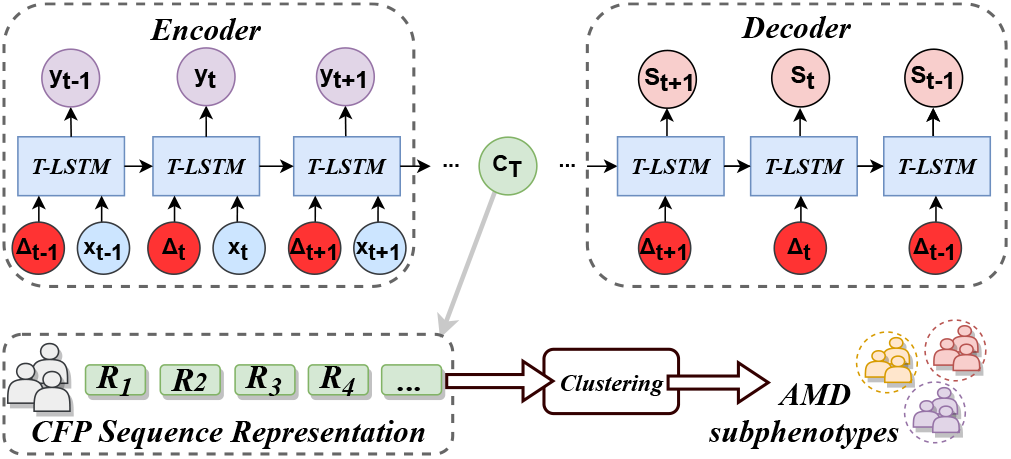
**We use an auto-encoder represent the CFP sequences as vectors. Encoder takes visits’ feature** *x*_*t*_ **and time gaps** Δ_*t*_ **as inputs. Decoder output the previous AMD steps** *s*_*t*_ **at different time. The memory vector** *C*_*T*_ **contains the CFP sequence information of the eye. Finally, we group eye representations** *C*_*T*_ **into AMD subphenotypes**.

### 2.2 CFP Feature Extraction with Contrastive Attention Module

Given a CFP *v*_*t*_ in *t*^*th*^ visit, we adopt DenseNet [16] to extract the image feature maps:

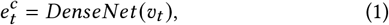

where 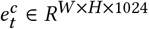 is the output feature map before the average pooling layer of DenseNet. An attention module is adopted to automatically focus on the abnormal area in eye images.

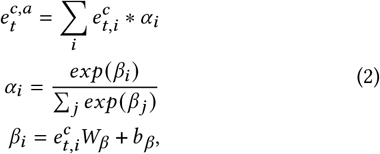

where 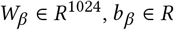 are learnable parameters. 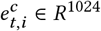 is the *i*^*th*^ vector of 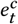.

#### Contrastive attention module

Due to the imbalanced late AMD distribution on various patient groups (e.g., with different ages, gender, smoking history), deep learning models might learn the bias and discrimination from the data. We present a contrastive attention module to make model be fair and focus on abnormalities. We divide all the healthy eyes or early AMD stage eyes in training set into various pools based on demographics (i.e., gender, age, smoking history). Given an eye image of a patient, we first collect the CFP feature vectors in the same pool as the patient, denoted as *P* = {*p*_1_, *p*_2_, …, *p* _*P*_ }, *p*_∗_ ∈ *R*^1024^. An aggregate attention is introduced to generate the weighted average feature vector 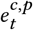 as thecommon feature vector of the patient pool, where the weight is set as the cosine similarity.

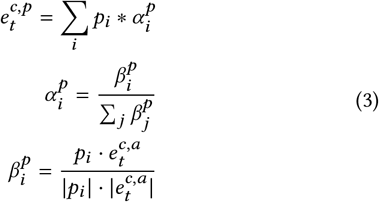

To obtain the contrastive information 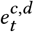, we remove (i.e., subtract) the common feature 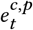 from the feature vector of the input image 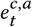.

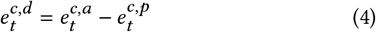

### 2.3 AMD Stage Classification

There are 12 steps (i.e., step 1-12) and 3 categories (i.e., early AMD, iAMD, late AMD) for AMD disease [10]. To make the CNN model learn more accurate and fine-grained features, our model predicts the probabilities for the 12 AMD steps. Intuitive loss function for the multi-class classification task is cross-entropy loss. However, the loss function fails to consider the semantic gap between AMD steps (e.g., the difference from step 4 to step 5 is much smaller than the difference from step 4 to step 9), which might be harmful for the feature extraction and future late AMD stage prediction. Thus we convert the 12-class classification task to 12 binary-class classification tasks. Our model predict the ground truth 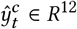 and 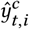 (i.e., the *i*^*th*^ dimension of 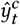) denotes whether the AMD step is higher than step *i* at time *t*. For example, given a CFP with AMD step equal to 3, we use 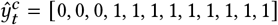 as the label vector.

Given CFP feature 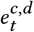, fully connected layers and Sigmoid layers are followed to generate the AMD probabilities for current visit:

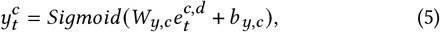

where *W*_*y,c*_ ∈ *R*^1024×12^, *b*_*y,c*_ ∈ *R*^12^ are learnable parameters. 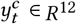 denotes the probability for 12 AMD steps at time *t*.

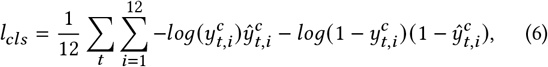

where 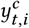 and 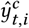 are the predicted probability and ground truth for AMD step *i* at time *t*. After the AMD stage classification model is well trained, we assume the CNN model can extract the abnormal features from CFPs.

### 2.4 Sociodemographic and AMD Progression Embedding

#### Sociodemographic embedding

Following [3], we extract patients’ sex, age, race, body mass index and smoking history as sociodemographic information and represent them as binary vectors. Given the sociodemographic vector *d*_*t*_ 0, 1 ^*m*^ at *t*^*th*^ step. We use fully connected layers to map *d*_*t*_ to sociodemographic feature vector 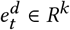.

#### Genotypic information embedding

Following [19, 23], we incorporate 52 AMD-associated independent genetic variants reported by the International AMD Genomics Consortium [13] to the patient representation module. We map 52 AMD genetic risk score to vector *e*^*g*^ with fully connected layers.

#### Progression information embedding

Following [3], we also utilize patients’ previous AMD category information in late AMD prediction. Different from [3] that just concatenates the last visit’s AMD category and demographic vectors, we present two kinds of progression information embedding methods to map previous AMD category sequences to embedding vectors.

In the first method, we assume all the previous AMD categories have been correctly identified by clinicians. We represent the AMD progression information during the previous visits as a vector 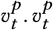 consists of two kinds of information, as shown in Figure 2 (D). The first is the AMD stage history during the last years’ visits. We sample the AMD stage for every half year. Patients might miss some routine follow-ups and the AMD stage vectors are not fully observed. We impute the missing AMD stages with linear interpolation. The second kind of information of 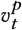 is the number of years that it takes for the individual eye to progress from step *i* to step *j*, where 1 ≤ *i* < *j* ≤ 9. We concatenate the two vectors to generate 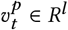. In our implementation, we set the stage observation window as 6 years. Then we use a fully connected layer to map the progression vector 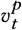 to an embedding vector 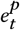 :

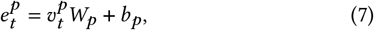

where *W*_*p*_ ∈ *R*^*l*×*k*^, *b*_*p*_ ∈ *R*^*k*^ are learnable parameters. 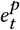 denotes the AMD progression embedding, which contains the information how the AMD disease of the patient’s eye progress during the last years’ visits. We concatenate the multi-modal features 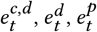 and *e*^*g*^ and adopt a fully connected layer to produce *x*_*t*_, which is sent to a time-aware LSTM to model the eye’s health states.

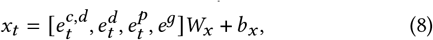

where *W*_*x*_ ∈ *R*^4*k*×*k*^ and *b*_*x*_ ∈ *R*^*k*^ are learnable variables. [·, ·, ·, ·] denotes the concatenation operation.

In the second method, we use the stage prediction results 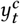 in Eq. (5) to generate progression embedding vector. The progression embedding method requires manual work for grading the AMD images. To develop an automatic late AMD prediction framework, we use the stage prediction results rather than the ground truth of previous AMD stages. Similarly, we utilize the predicted AMD categories during the last 6 years and conduct linear interpolation to impute the missing visits. Then we obtain a matrix *Y*_*t*_ ∈ *R*^12×12^ to represent the previous AMD progression information. Then we adopt 1-D convolutional layers to map the vector sequences to produce progression vector 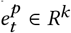.

### 2.5 Late AMD Stage Prediction with Time-aware LSTM

To capture the temporal patterns of AMD disease progression, we utilize LSTM [15] to model the CFP feature sequences. To address the irregular time gaps between visits, following [2], we introduce time-aware LSTM, which adjust the memory vector *C*_*t*−1_ to 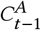 based on the time gap Δ_*t*_ as follows:

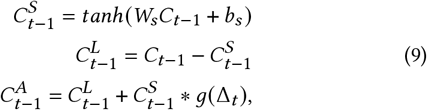

where *W*_*s*_ ∈ *R*^*k*×*k*^ and *b*_*s*_ ∈ *R*^*k*^ are learnable parameters. We first divide the memory vector *C*_*t*−1_ into long-term memory 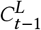 and short-term memory 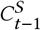. The sum of long-term memory and discounted short-term memory is used as the adjusted memory vector 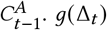 serves as the discount function and we use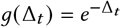 in our experiments. Given the adjust memory vector, we compute the new memory cell *C*_*t*_ and hidden state *h*_*t*_ :

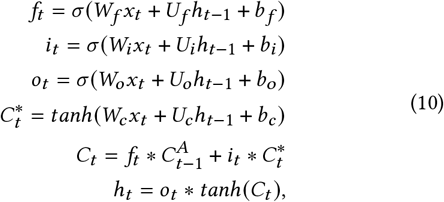

where *W*_*f*_, *W*_*i*_, *W*_*o*_, *W*_*c*_ ∈ *R*^*k*×*k*^, *U*_*f*_, *U*_*i*_, *U*_*o*_, *U*_*c*_ ∈ *R*^*k*×*k*^ and *b*_*f*_, *b*_*i*_, *b*_*o*_, *b*_*c*_ ∈ *R*^*k*^ are learnable parameters. Fully connected layers and Sigmoid layers are followed to generate the late AMD probabilities:

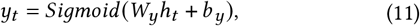

where *W*_*y*_ ∈ *R*^*k*^, *b*_*y*_ ∈ *R* are learnable parameters.

We use binary cross entropy to train the time-aware LSTM.

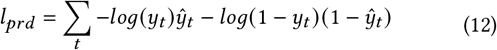

### 2.6 AMD Subtyping with Auto-encoder

To further study the AMD progression patterns among different patients’ eyes, we subtype individual eyes based on their temporal CFP sequences. We first project the varying-length CFP sequences to vectors with CAT-LSTM.

As shown in Figure 3, the T-LSTM in Eq. (10) serves as the encoder to extract the temporal sequence information. We build another T-LSTM as the decoder to reconstruct the stage history of the previous CFP images. The hidden state and the cell memory of the T-LSTM encoder at the end of input sequence are used as the initial hidden state and the memory content of the T-LSTM decoder. The first input time gap of the decoder is set to zero and it outputs the AMD stage in the last visit. Then the time gaps between visits are sent to T-LSTM decoder. When the reconstruction error is minimized, T-LSTM encoder learns the effective representations of CFPsequences.

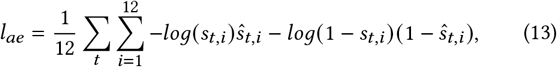

where *s*_*t,i*_ and *ŝ*_*t,i*_ are the predicted probability and ground truth for AMD step *i* at time *t*. Note that *s*_*t*_ and *ŝ*_*t*_ have the same format as 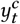 and 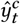 in Eq. (6).

The auto-encoder and CAT-LSTM are jointly trained when subtyping CFP sequences. We use a hyper-parameter *λ* (0 *< λ <* 1) to adjust the weights of the two loss functions.

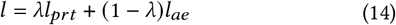

Note that we only jointly train the late AMD prediction model and auto-encoder when subtyping CFP sequences. When conducting late AMD prediction experiments, we use the binary cross entropy loss in Eq. (12) to train CAT-LSTM.

## 3 EXPERIMENT

To demonstrate the effectiveness of the proposed model, we conduct experiments on a real-world dataset and compare the proposed model with the-state-of-art methods.

### 3.1 Datasets and Settings

The AREDS is a multi-center prospective cohort study of the clinical course, prognosis, and risk factors of AMD [21]. 4,612 participants aged 55–80 years are recruited from 1992 at 11 retinal specialty clinics in the United States. The inclusion criteria are wide, from no AMD in either eye to late AMD in one eye. The AREDS data set is publicly accessible to researchers by request at dbGAP^2^. The statistics of the extracted data are displayed in Table 1. Note that in a visit, there might be multiple CFPs for individual eyes (e.g., from left and right sides). We randomly select one CFP in the training process, and use the average features from multiple images in the test phase.

**Table 1:**
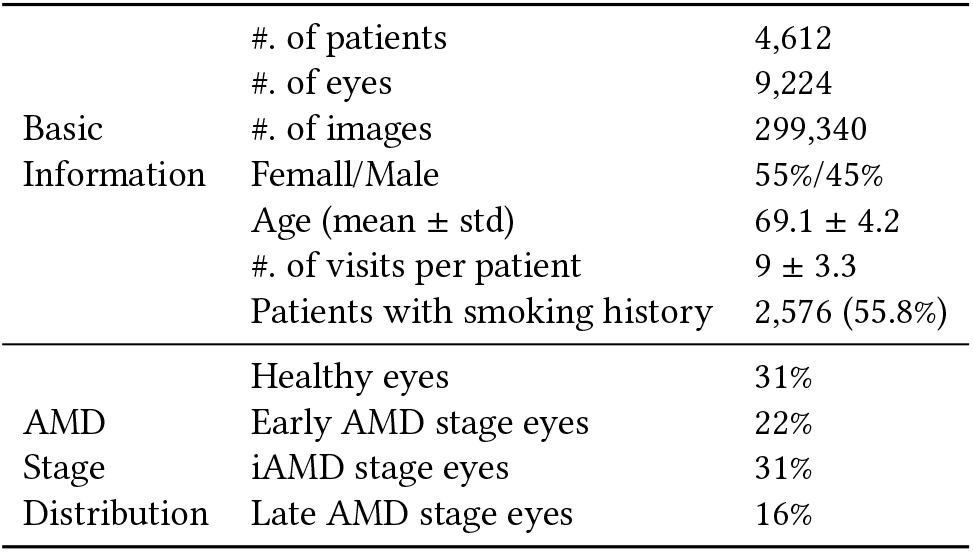
Statistics of AREDS.

We conduct experiments on two kinds of settings. The first is late AMD detection: to detect whether the patients’ eyes have progressed to late AMD stage. The second setting is late AMD prediction. Following [19, 23], we predict whether patients’ eyes will progress to late AMD within *n* years. Figure 4 shows the definitions of positive and negative samples. We conduct experiments with different n (i.e., n=1,2,3,4,5 and All). For patients who progress to late AMD at time *t*_*l*_, the visits between time *t*_*l*_ −*n* to *t*_*l*_ are set as positive samples, while the visits before time *t*_*l*_ − *n* are set as negative samples. When n is All, we set all the visits before *t*_*l*_ as positive samples. For patients who do not progress to late AMD ultimately, *t*_*e*_ is the time of last visit and all the visits before time *t*_*e*_− *n* are set as negative samples. When n is All, we set all the visits before time *t*_*e*_ as negative samples. The numbers of positive and negative samples in various settings can be found in supplementary materials (Table 9). Note that We remove the patients with less than 4 visits and the patients’ eyes with late AMD in the first visit when building the dataset.

**Figure 4:**
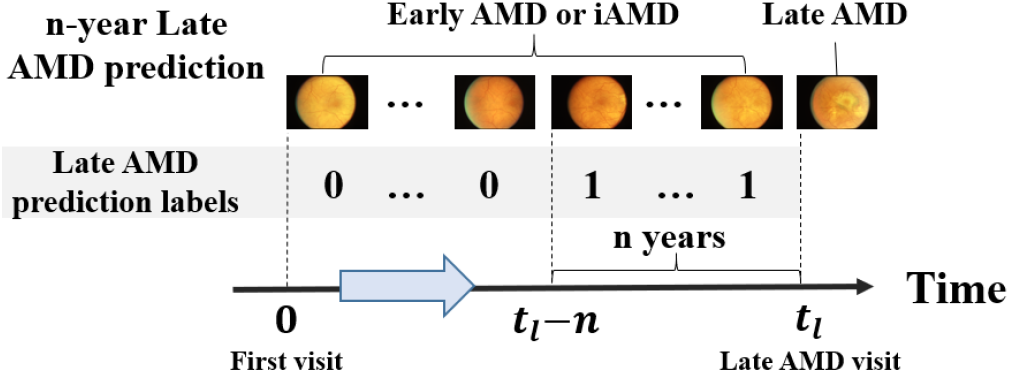
**Settings of n-year late AMD prediction. We predict whether patients’ eyes will progress to late AMD within n years**. *t*_*l*_ **is the time of the first late AMD visit. The visits at time** *t* (*t*_*l*_ − *n* ≤ *t < t*_*l*_) **are set as positive samples. The visits before** *t*_*l*_ − *n* **are set as negative samples**.

### 3.2 Methods for Comparison

We compare the proposed model with late AMD detection and prediction methods:

- **DeepSeeNet** [18]: DeepSeeNet is developed to automatically classify patients by the AREDS Simplified Severity Scale (score 0-5) using bilateral CFP. DeepSeeNet also has an output branch to generate the risk of late AMD stage. We modify the output branch to predict AMD progression within coming years.
- **Chen et al**. [7]: Chen et al. detect four AMD characteristics (drusen area, geographic atrophy, increased pigment, and depigmentation), then combine them to derive the overall 9-step score. We add an output branch to predict the risk of late AMD stage.
- **Babenko et al**. [1]: Babenko et al. adopt Inception-v3 to predict AMD progression for a stereo pair of left and right sides of the same eye. The averaged risk is used as the final output.
- **Yan et al**. [23]: Yan et al. utilize Inception-v3 to extract deep image features. The deep features with 52 independent genetic variants are fed to another fully connected layer to predict the time to late AMD development exceeding certain inquired years.
- **Peng et al**. [19]: Peng et al. combine the deep features generated by DeepSeeNet and genotypic information to represent the patients’ eyes, then adopt a survival model to predict AMD progression risk.
- **Bhuiyan et al**. [3]: Bhuiyan et al. use a two-step ensemble method to predict AMD progression. First, a screening module classifies the images into 12-class severity scales with five deep learning frameworks (e.g., Inception-V3, Xception, Inception-Resnet-v2). Then the resulting AMD scores are combined with sociodemographic clinical data (including age, race, sex, body mass index, visual acuity, and sunlight exposure) and other automatically extracted imaging data by a logistic model tree machine learning technique to predict risk for progression to late AMD.

#### The proposed methods

We developed two versions of CAT-LSTM with different progression embeddings:

- **CAT-LSTM-v1**: We send the output of CA-CNN to T-LSTM to generate the risk of progression to late AMD stage. We use the ground truth of previous AMD progression information in this version.
- **CAT-LSTM-v2**: To build an automatic AMD prediction method, we use the AMD stage probability produced by CA-CNN rather than the manual labeled AMD stage to generate the progression information.

### 3.3 Implementation Details

We implement our proposed CAT-LSTM models with PyTorch 0.4.1^3^. For training models, we use Adam optimizer with a mini-batch of 8 patients. For each patient, we send the CFP images in recent 8 visits to our model. We train on 4 GPUs (TITAN RTX 2080) for 40 epochs, with a learning rate of 0.0001. We randomly divide the patients in the dataset into 10 sets. All the experiment results are averaged from 10-fold cross validation, in which 7 sets are used for training every time, 1 set for validation and 2 sets for test. The validation sets are used to determine the best values of parameters in the training iterations. We first pretrain CA-CNN with loss *l*_*cls*_ in Eq. (6) and then train the whole CAT-LSTM model with loss *l*_*prd*_ in Eq. (12). We use the area under the receiver operating characteristic curve (AUROC) in the test sets as a measure for comparing the performance of all the methods. The CFP images, sociodemographics, genotype and progression information are projected into 1024-d vectors. We set the hidden state of T-LSTM as 256-d vectors. More details can be found at GitHub^1^.

### 3.4 Late AMD Detection and Prediction

Table 2 displays the late AMD detection and prediction results. The results show that all the methods achieved similar performance on the late AMD detection task. The reason is that when an eye progress to late AMD stage, the eye images would look very different from healthy eyes (e.g., large drusen area or pigmentary abnormalities), as shown in Figure 1. Deep learning models can easily capture the abnormalities and accurately classify the images. In this subsection, we mainly discuss the late AMD prediction tasks, which are more significant for timely clinical intervention for highrisk patients. The proposed model outperforms the baselines on the late AMD prediction tasks by more than 3% on AUROC, which demonstrates the effectiveness of our model.

**Table 2:** **AUROC of late AMD stage prediction. Late AMD detection denotes detecting whether the patients’ AMD diseases have progressed to late stages based on information up to current visit. Late AMD prediction (1-5 years and All-year) denote predicting whether patients’ AMD diseases will progress to late AMD within 1-5 years and progress to late AMD ultimately**.

| Methods | Late AMD Detection | n-year Late AMD Prediction |  |  |  |  |  |
| --- | --- | --- | --- | --- | --- | --- | --- |
|  |  | 1-year | 2-year | 3-year | 4-year | 5-year | All-year |
| DeepSeeNet [18] | 0.992($\pm 0.003$) | 0.873($\pm 0.011$) | 0.873($\pm 0.010$) | 0.875($\pm 0.011$) | 0.878($\pm 0.013$) | 0.875($\pm 0.012$) | 0.875($\pm 0.013$) |
| Chen et al. [7] | 0.993($\pm 0.003$) | 0.875($\pm 0.010$) | 0.874($\pm 0.012$) | 0.880($\pm 0.010$) | 0.878($\pm 0.012$) | 0.879($\pm 0.013$) | 0.877($\pm 0.014$) |
| Bhuiyan et al. [3] | 0.993($\pm 0.004$) | 0.872($\pm 0.011$) | 0.874($\pm 0.010$) | 0.878($\pm 0.012$) | 0.875($\pm 0.013$) | 0.877($\pm 0.011$) | 0.877($\pm 0.012$) |
| Babenko et al. [1] | **0.994($\pm 0.002$)** | 0.879($\pm 0.012$) | 0.880($\pm 0.012$) | 0.884($\pm 0.011$) | 0.882($\pm 0.014$) | 0.884($\pm 0.013$) | 0.883($\pm 0.012$) |
| Yan et al. [23] | **0.994($\pm 0.003$)** | 0.887($\pm 0.013$) | 0.890($\pm 0.013$) | 0.895($\pm 0.012$) | 0.894($\pm 0.013$) | 0.895($\pm 0.013$) | 0.895($\pm 0.013$) |
| Peng et al. [19] | **0.994($\pm 0.002$)** | 0.886($\pm 0.014$) | 0.891($\pm 0.013$) | 0.894($\pm 0.012$) | 0.894($\pm 0.014$) | 0.895($\pm 0.014$) | 0.895($\pm 0.013$) |
| CAT-LSTM-v1 | **0.994($\pm 0.003$)** | **0.925($\pm 0.009$)** | **0.924($\pm 0.012$)** | **0.925($\pm 0.010$)** | **0.924($\pm 0.011$)** | **0.925($\pm 0.010$)** | **0.925($\pm 0.010$)** |
| CAT-LSTM-v2 | **0.994($\pm 0.002$)** | **0.905($\pm 0.012$)** | **0.907($\pm 0.010$)** | **0.907($\pm 0.011$)** | **0.906($\pm 0.010$)** | **0.907($\pm 0.010$)** | **0.907($\pm 0.011$)** |

Among the baseline methods, with the genotype information, Yan et al. [23] and Peng et al. [19] outperform the other models, the reason is that the two methods incorporate more information from the genetic risk scores of 52 independent genetic markers. Babenko et al. [1] performs better than DeepSeeNet [18] Chen et al. [7] and Bhuiyan et al. [3], we speculate the reason is that the model takes both left and right sides of same eyes as input and thus can capture the global features of the eyes. Both versions of our model in Table 2 outperform the baselines, which can demonstrate the effectiveness of our model. With additional ground truth information of AMD stage in previous visits, CAT-LSTM-v1 outperforms the baselines by more than 3% on AUROC. To fairly compare with the baselines, CAT-LSTM-v2 takes the predicted AMD stage by our model as inputs, and still outperforms the baselines.

Table 2 shows that each model achieves similar superior performance in different settings of late AMD prediction, while shortterm prediction is supposed to be easier than long-term prediction. We speculate the reason might be related to imbalanced positive/negative distribution is different settings. As Table 9 in supplementary materials shows, the numbers of positive samples in short-term late AMD prediction settings (e.g., *<* 2% positive samples in 1-year late AMD prediction setting) are much less than long-term AMD prediction settings (e.g., *>* 15% positive samples in 5-year late AMD prediction setting), so the short-term prediction models cannot outperform the long-term prediction models with much less positive samples. Our prediction results also align with existing late AMD prediction studies [19, 23].

#### Ablation study

To further investigate the contribution of each component (i.e., contrastive attention module, AMD progression embedding, genotypic information embedding) of our model, we conduct ablation study by comparing additional four versions of the proposed model:

- **CNN**: We only use a CNN model (i.e., DenseNet) to predict the risk of progression to late AMD stage after 5 years.
- **CA-CNN**: To demonstrate the effectiveness of the contrastive attention module, we use the CNN model with contrastive attention module to predict the risk of progression to late AMD stage after 5 years.
- **CA-CNN+Genotype**: To demonstrate the effectiveness of genotype information, we incorporate the genotype embedding vector when representing patient health states and predict the risk of progression to late AMD stage after 5 years.
- **CAT-LSTM**^−*p*^ : To demonstrate the effectiveness of progression information, we remove the progression embedding and only use CFP images, genotype and sociodemographics information to predict AMD progression.

Since area under Precision-Recall curves (AUPRC) give a more informative picture of an algorithm’s performance than AUROC in imbalanced datasets [9], we add AUPRC to evaluate the various versions of our model. Table 3 displays the prediction results on AUROC and AUPRC. CA-CNN outperforms CNN by more than 2% on AUPRC, which demonstrates the effectiveness of contrastive attention module. We speculate the reasons are two-fold: (i) contrastive attention module force the model to focus on the difference between the input image and healthy eye images, which might be the abnormal regions of the input image; (ii) the late AMD stage rates among different patient groups vary a lot as shown in Table 8, contrastive attention module remove the common information in patient groups, which make the feature representation fairer and thus can improve the overall prediction performance. With the consideration of genotypic information, CA-CNN+Genotype outperforms CA-CNN on both AUROC and AUPRC, which demonstrates that genotype data could improve the AMD progression prediction performance. CAT-LSTM^−*p*^ outperforms the CA-CNN+Genotype, which demonstrates that with previous visits’ information, T-LSTM can improve the prediction results by capturing the AMD stage progression information. Inspired by the thought, we directly embed the progression information and input to our model in CAT-LSTM-v1 and CAT-LSTM-v2. With the ground truth of AMD stages in previous visits, CAT-LSTM-v1 outperform the other versions a lot on both AUROC and AUPRC. Even without manually labeled AMD stage as inputs, CAT-LSTM-v2 still performs much better than other versions, which further demonstrates capturing previous AMD progression information does help the prediction for future AMD progression.

**Table 3:** AUROC and AUPRC of late AMD prediction.

| Methods | 5-year AMD prediction |  |
| --- | --- | --- |
|  | AUROC | AUPRC |
| CNN | 0.882($\pm 0.011$) | 0.416($\pm 0.031$) |
| CA-CNN | 0.891($\pm 0.011$) | 0.441($\pm 0.032$) |
| CA-CNN + Genotype | 0.897($\pm 0.012$) | 0.458($\pm 0.033$) |
| CAT-LSTM<sup>-P</sup> | 0.899($\pm 0.011$) | 0.467($\pm 0.034$) |
| CAT-LSTM-v1 | 0.925($\pm 0.010$) | 0.557($\pm 0.035$) |
| CAT-LSTM-v2 | 0.907($\pm 0.010$) | 0.512($\pm 0.034$) |

### 3.5 AMD Subtyping

We cluster the patients’ eyes based on their fundus image sequences. The auto-encoder can output the previous AMD stages during the last years’ visits. We can assume the hidden state vector input to the decoder contains the progression information and can represent the whole fundus image sequence. We cluster the hidden state vector with k-means and obtain 3 subphenotypes. Note that we only use patients’ first 4 years’ visits of early and intermediate AMD stages for clustering.

#### Subphenotype characteristics

The cluster descriptive statistics of AMD subphenotypes are shown in Table 4. The subphenotype I has the most individual eyes, youngest age and the lowest AMD stage rate. The subphenotype III has the least individual eyes, but more than half the eyes in the subphenotype would progress to late AMD stages. We find that age and smoking history have positive correlations with the late AMD stage rate. Moreover, subphenotype III has the maximal abnormal rates (e.g., depigmentation, increased pigment and drusen abnormalities) than the other two subphenotypes. Besides, we compute the progression time from earlier AMD steps to later AMD steps and find the disease progression for patients’ eyes in subphenotype III is faster than the AMD progression in the other two subphenotypes, which further demonstrates the AMD progression speed in previous years has correlation with the probability of progressing to late AMD stage in coming years. It can also explain why the proposed progression embedding and LSTM model can improve the late AMD prediction performance.

**Table 4:**
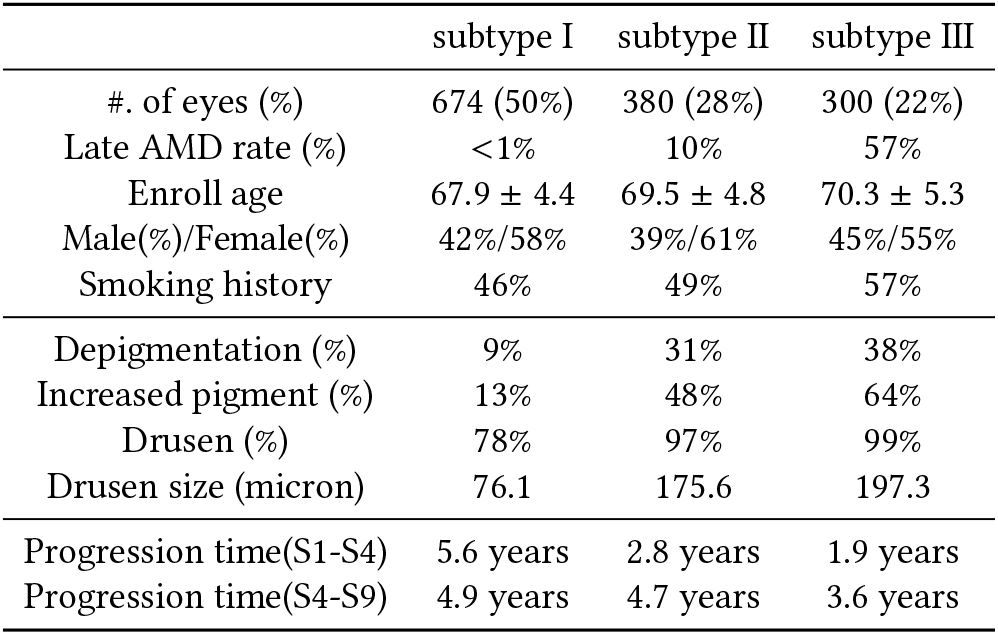
**Cluster descriptive statistics of AMD subphenotypes. We group patients into 3 subphenotypes based on their fundus images and sociodemographics up to intermediate AMD stage. Progression time denote the average number of years for patients with AMD to progress from earlier steps (i.e**., **step 1 and step 4) to later steps (i.e**., **step 4 and step 9)**.

Existing study [13] has reported that 52 independent genetic variants are associated with AMD disease. We compute the distribution of the genetic variants across 3 subphenotypes. Figure 7 displays the genetic variants with significant distribution difference (P-value *<* 0.05) across the 3 subphenotypes. Because the alternative allele rates of different genetic markers vary a lot, we normalize the rates when visualizing the distribution of the genetic variants. Based on the different characteristics (e.g, sociodemographics, abnormalities, AMD progression speed in Table 4, and genetic variant distribution in Figure 7), we might be able to early identify patients’ AMD subphenotype and provide timely medical interventions for patients with high risk of progression to late AMD stage, which might improve the treatment effects or alleviate the disease progression.

#### Clustering evaluation

To evaluate the clustering performance, we compare the proposed CAT-LSTM with a baseline CNN+LSTM, which adopts CNN to represent CFP as feature vectors and leverage LSTM to extract CFP sequence features. We use the same k-means method to group the CFP sequences for CNN+LSTM. Table 5 displays the clustering results. Since we do not know the ground truth AMD subphenotypes, we cannot measure the clustering performance with common clustering evaluation metrics such as purity and rand index. Instead, we use two popular metrics CalinskiHarabasz Index (CHI) [6] and Davis-Bouldin Index (DBI) [8], which can measure the performance of clustering algorithms on label-unknown dataset. Note that CHI is related to the size of the dataset, we normalize the value by dividing CHI by the number of patients. The results show that both versions of CAT-LSTM perform better than CNN+LSTM on both metrics.

**Table 5:**
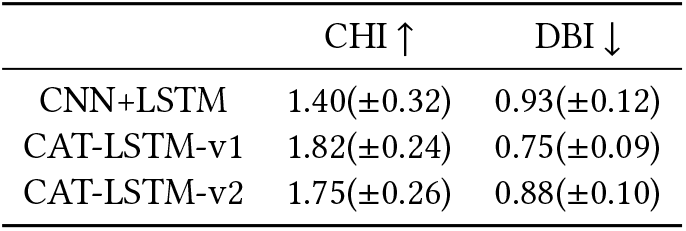
**Clustering performance. CAT-LSTM denotes our CAT-LSTM model jointly trained with auto-encoder. Note that higher CHI and lower DBI relate to a model with better separation between the clusters**.

| | CHI ↑ | DBI ↓ |
| --- | --- | --- |
| CNN+LSTM | 1.40($\pm 0.32$) | 0.93($\pm 0.12$) |
| CAT-LSTM-v1 | 1.82($\pm 0.24$) | 0.75($\pm 0.09$) |
| CAT-LSTM-v2 | 1.75($\pm 0.26$) | 0.88($\pm 0.10$) |

#### Visualization of learned CFP sequence representations

We adopt the t-distributed stochastic neighbor embedding (t-SNE) algorithm [17] to project all CFP sequences of individual eyes into a 2D space and Figure 5 shows the visualization results. To fairly compare with CNN+LSTM, we use CAT-LSTM-v2 (which embeds progression features based on predicted AMD stage rather than ground truth) to extract CFP sequence features. Based on which stage the individual eye progress to ultimately, we divide the eyes into 3 groups (i.e., early AMD stage, iAMD stage and late AMD stage) with different colors in Figure 5. Note that we just use the CFP sequences up to iAMD stage for Figure 5 (a) and Figure 5 (b), and use all the CFP sequences (including late AMD stage) for Figure 5 (c). As Figure 5 (a) shown, CNN+LSTM cannot distinguish the iAMD and late AMD groups well at early time. With the AMD progression information and T-LSTM to model the CFP sequences, CAT-LSTM performs better on identifying different groups as Figure 5 (b) shown. Given the whole CFP sequences (including CFPs in late AMD stages) as input, our model can clearly identify the 3 groups as Figure 5 (c) shown.

**Figure 5:**
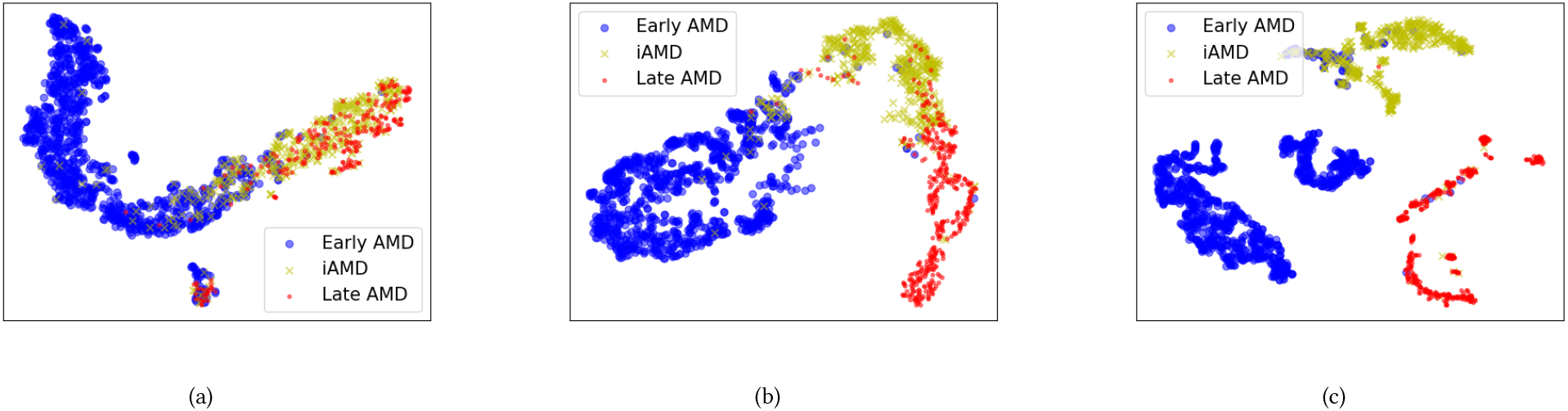
**Projection scatter plot of individual eyes based on features extracted by (a) CNN+LSTM, (b) CAT-LSTM (with CFP inputs up to iAMD stage) and (c) CAT-LSTM (with CFP inputs including late AMD stages). To fairly compare with CNN+LSTM, we use CAT-LSTM-v2 (which embeds progression features based on predicted AMD stage rather than ground truth) to extrace CFP sequence features**.

## 4 RELATED WORK

In this section, we briefly review the existing works related to late AMD detection and prediction.

### AMD Detection

Burlina et al. [5] compare the performance of humans and deep learning in grading CFP to detect AMD. Peng et al. [18] present DeepSeeNet to classify patients automatically by the AREDS Simplified Severity Scale (score 0-5) using bilateral CFP. Grassmann et al. [14] utilize an ensemble of neural network architectures to classify CFPs into 12-step categories. Burlina et al. [4] adopt deep convolutional neural networks to predict the AREDS 9-step detailed severity scale for AMD to estimate 5-year risk probability with reasonable accuracy. Chen et al. [7] detect four AMD characteristics (drusen area, geographic atrophy, increased pigment, and depigmentation), then combine them to derive the overall 9-step score.

### AMD prediction

Babenko et al. [1] adopt Inception-v3 to predict AMD progression for stereo pairs of different sides of the same eye. Yan et al. [23] utilize Inception-v3 to extract deep image features. The deep features with 52 independent genetic variants are fed to another fully connected layer to predict the time to late AMD development exceeding certain inquired years. Peng et al. [19] combine the deep features generated by DeepSeeNet and genotypic information to represent the patients’ eyes, then adopt a survival model to predict AMD progression risk. Bhuiyan et al. [3] propose to ensemble five deep learning frameworks (e.g., Inception-V3, Xception, Inception-Resnet-v2) to predict AMD progression. The resulting AMD scores of various models are combined with sociodemographic clinical data (including age, race, sex, body mass index, visual acuity, and sunlight exposure) and other automatically extracted imaging data by a logistic model tree machine learning technique to predict risks of progressing to late AMD.

Although the methods described above have achieved superior performance on AMD detection and prediction, they do not make full use of the CFP sequence data and AMD progression information in late AMD prediction tasks, which might limit their prediction performance. In this study, we present progression embedding and introduce T-LSTM to capture AMD progression information, which significantly improves the prediction performance.

## 5 CONCLUSION

We proposed a new AMD progression prediction framework CAT-LSTM. The framework adopts CNN to extract fundus image features and a time-aware LSTM to model CFP sequence feature, sociodemographics, genotype and AMD progression information. We introduce a contrastive attention module to force the framework to focus on the abnormal area of images. To explicitly utilize the AMD progression information during the last years’ visits, we present a progression embedding module to map the AMD step sequences to a vector. Experiments on real-world datasets have shown that all the contrastive attention modules, progression embedding, and T-LSTM can improve late AMD progression performance. Moreover, we represent patients’ temporal image sequences as fixed-size vectors with an auto-encoder and subtype the CFP sequences with k-means based on the learned representation. The subtyping analysis shows that the patients in the 3 subphenotypes have different probabilities of progressing to the late AMD stage. The proposed AMD subtyping framework is useful in identifying patients with a high risk of progressing to the late AMD stage in patients’ iAMD stage, which paves the way for improved personalization of AMD management.

## Data Availability

All data in the study are available online at

https://www.ncbi.nlm.nih.%20gov/projects/gap/cgi-bin/study.cgi?study_id=phs000001.v3.p1

## A IMPORTANT NOTATIONS

We list the important notations in Table 6.

**Table 6:** Important notations.

| Notation | Definition |
| --- | --- |
| $T$ | The number of visit |
| $V$ | The CFP sequence |
| $g$ | The genotype vector |
| $e^g$ | The embedding vector of $g$ |
| $v_t$ | The CFP image in the $t^{th}$ visit |
| $e_t^{c,a}$ | The CFP feature generated by attention module |
| $p_i$ | The CFP feature vector in patient pool |
| $e_t^{c,p}$ | The aggregate features |
| $e_t^{c,d}$ | The abnormal features generated by CA |
| $D$ | The sociodemographic sequence |
| $d_t$ | The sociodemographic vector in the $t^{th}$ visit |
| $e_t^d$ | The embedding vector of $d_t$ |
| $v_t^p$ | The AMD progression vector before the $t^{th}$ visit |
| $e_t^p$ | The embedding vector of $v_t^p$ |
| $y_t^c$ | The predicted AMD stage probability in the $t^{th}$ visit |
| $\hat{y}_t^c$ | The ground truth of AMD stage in the $t^{th}$ visit |
| $y_t$ | The predicted late AMD probability after $t^{th}$ visit |
| $\hat{y}_t$ | The late AMD ground truth after $t^{th}$ visit |
| $W_*, b_*$ | The learnable parameters |
| $\Delta_t$ | The elapsed time between $t - 1^{th}$ and $t^{th}$ visit |
| $x_t$ | The combination of multi-modal features in $t^{th}$ visit |
| $C_t$ | The memory vector of T-LSTM in $t^{th}$ visit |
| $h_t$ | The hidden state of T-LSTM in $t^{th}$ visit |

## B HYPER-PARAMETER OPTIMIZATION

There is a hyper-parameter *λ* in Eq. (14). Note that we only jointly train the late AMD prediction model and auto-encoder when subtyping CFP sequences. We use the clustering evaluation metrics CHI and DBI to select the value of *λ*. As Table 7 shown, when 0.3 ≤ *λ* ≤ 0.9, clustering performance is not sensitive to *λ*. In our experiment, we set *λ* = 0.5.

**Table 7:**
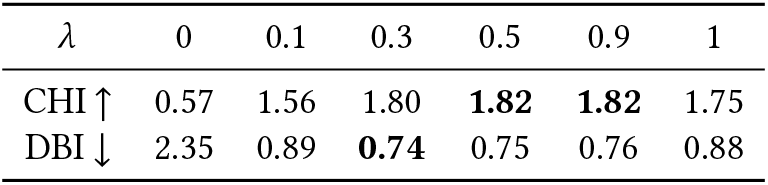
Hyper-parameter optimization for *λ* in Eq. (14).

| $\lambda$ | 0 | 0.1 | 0.3 | 0.5 | 0.9 | 1 |
| --- | --- | --- | --- | --- | --- | --- |
| CHI ↑ | 0.57 | 1.56 | 1.80 | **1.82** | **1.82** | 1.75 |
| DBI ↓ | 2.35 | 0.89 | **0.74** | 0.75 | 0.76 | 0.88 |

## C K SELECTION FOR K-MEANS

We try to use different K for k-means when clustering the CFP sequences. As shown in Figure 6, when *K* = 3, we have the best DBI value for CFP clustering. It is also the elbow point for CHI. Thus we cluster the CFP sequences into 3 subphenotypes.

**Figure 6:**
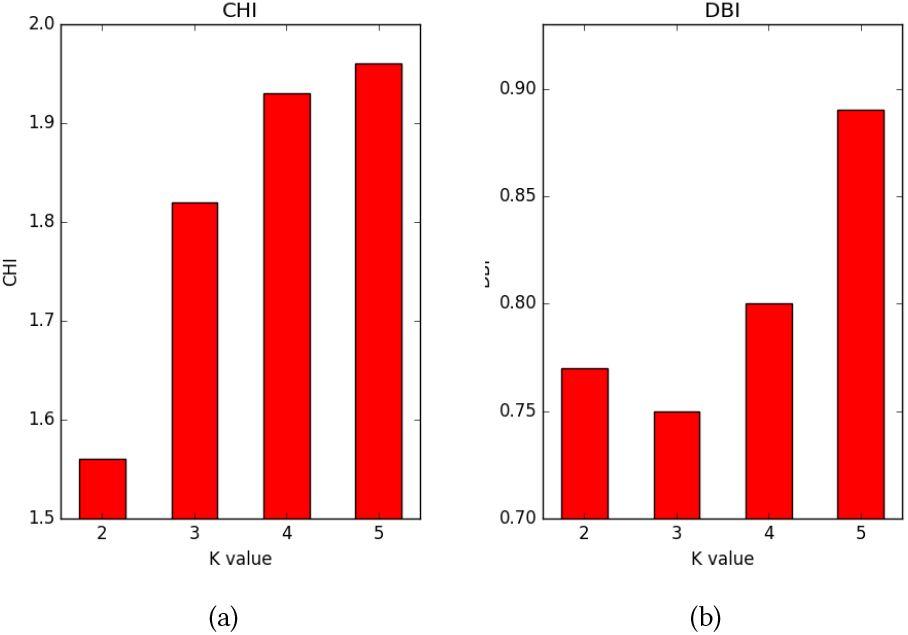
**CHI and DBI across different** *K* **for k-means to cluster the CFP sequences. When** *K* = 3, **we have the best DBI value for CFP clustering. It is also the elbow point for CHI**.

## D LATE AMD RATE IN VARIOUS PATIENT GROUPS

Patients with different demographics (e.g., gender and age) have different risks of progressing to late AMD stages. Table 8 displays the late AMD rates in various patients groups. Higher age (e.g., age *>* 80) and smoking history have positive correlations with late AMD rates. Due to the existence of the imbalanced late AMD distribution, machine learning methods might learn the bias and discrimination from the data. Thus we propose to use contrastive attention to remove the common features in patient groups and force the model to learn to focus on the abnormalities on fundus images.

**Table 8:** Late AMD rate in different patient groups.

| Sociodemographic | Value | Late AMD rate |
| --- | --- | --- |
| Gender | Female | 15.0% |
|  | Male | 13.8% |
| Smoking history | No | 13.1% |
|  | Yes | 16.7% |
| Age | < 70 | 11.2% |
|  | 70-80 | 14.7% |
|  | > 80 | 19.8% |

## E POSITIVE/NEGATIVE SAMPLE DISTRIBUTION

Table 9 displays the positive/negative sample distribution for late AMD detection and prediction settings.

**Table 9:** Positive/Negative sample distribution for late AMD prediction.

| Methods | Late AMD Detection | n-year Late AMD Prediction |  |  |  |  |  |
| --- | --- | --- | --- | --- | --- | --- | --- |
|  |  | 1-year | 2-year | 3-year | 4-year | 5-year | All-year |
| #. of positive visit samples | 9,207 | 753 | 1,558 | 2,240 | 2,861 | 3,390 | 4,713 |
| #. of negative visit samples | 57,262 | 37,375 | 32,371 | 27,497 | 23,020 | 17,498 | 33,415 |
| Positive rate | 16.07% | 1.97% | 4.59% | 7.53% | 11.05% | 16.22% | 12.36% |

## F GENETIC MARKER VISUALIZATION

We further analyze the alternative allele distribution of 52 AMD-associated genetic markers across various subphenotypes. Figure 7 displays the alternative allele distribution of AMD-associated genetic markers with significant distribution difference (P-value < 0.05) across 3 subphenotypes.

**Figure 7:**
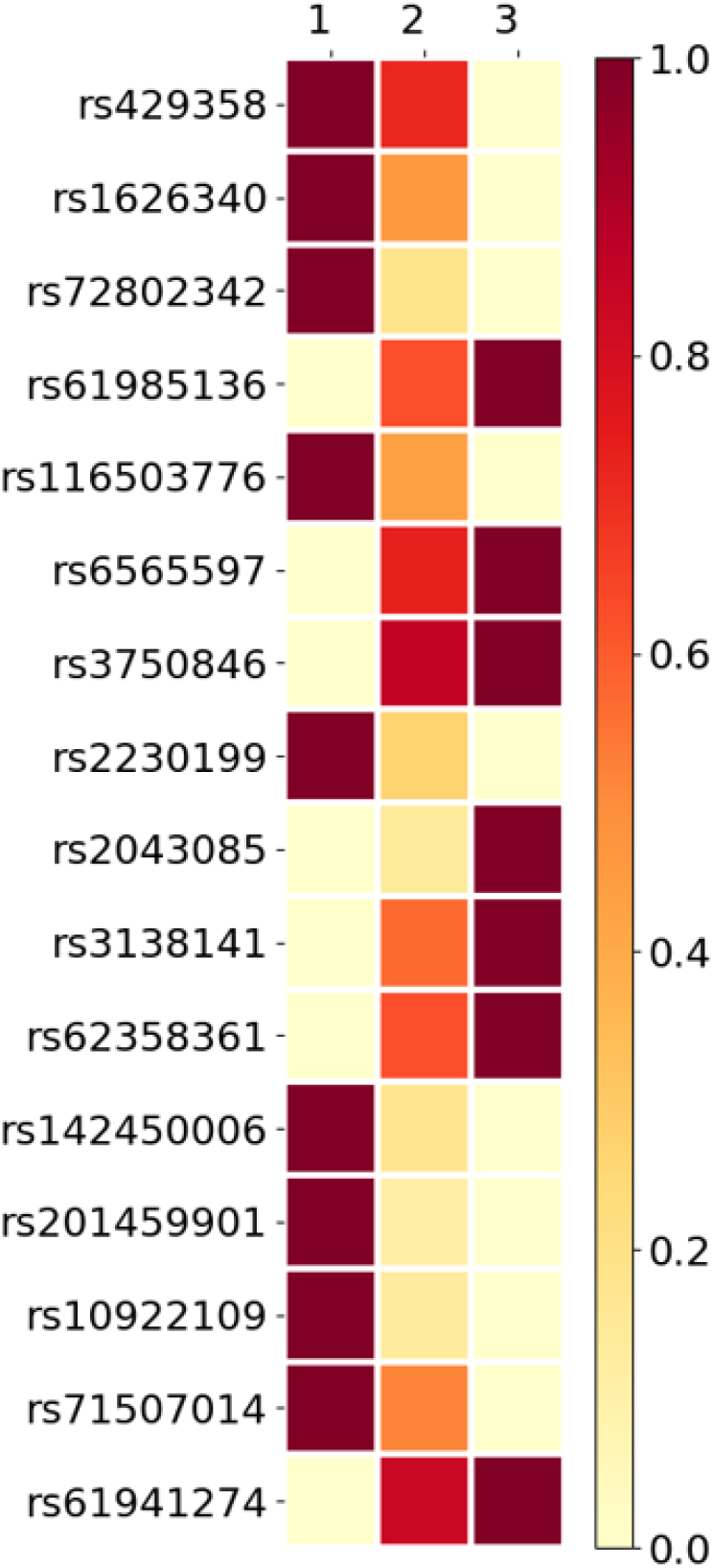
**The distribution of AMD-associated genetic markers’ alternative allele across the three subphenotypes. The subphenotypes are 1: subphenotype I; 2: subphenotype II; 3: subphenotype III. Because the alternative allele rates of different genetic markers vary a lot, we normalize the rates when visualizing the distribution**.

## Footnotes

1 https://github.com/yinchangchang/CAT-LSTM

2 https://www.ncbi.nlm.nih.gov/projects/gap/cgi-bin/study.cgi?study_id=phs000001.v3.p1

3 https://pytorch.org/

